# Genome- and Transcriptome-wide association meta-analysis reveals new insights into genes affecting coronary and peripheral artery disease

**DOI:** 10.1101/2025.01.27.25320452

**Authors:** Michael Rode, Maciej Rosolowski, Katrin Horn, Sylvia Henger, Andrej Teren, Kerstin Wirkner, Joachim Thiery, Markus Loeffler, Janne Pott, Holger Kirsten, Markus Scholz

## Abstract

**Background:** A low ankle-brachial Index (ABI) is an established condition for peripheral artery disease (PAD) and cardiovascular disease risk. The search for genetic determinants of the ankle-brachial index (ABI) is important to better understand molecular patho-mechanisms of PAD and its commonalities with cardiovascular diseases (CVD), supporting development of new drug targets and tailored preventive or therapeutic measures.

**Methods:** To search for genetic factors contributing to ankle-brachial index, we integrated genome-wide (GWAMA) and transcriptome-wide association analysis (TWAMA) of two German cohorts, the population-based LIFE-Adult cohort and LIFE-Heart, a cohort of patients with suspected or confirmed coronary artery disease. Pathway analysis of identified genes was used to explore biological mechanisms potentially involved in ABI pathophysiology. Finally, we analysed co-associations of known CAD or carotid plaque associations with ABI to detect possible genetic commonalities.

**Results:** By our GWAMA, we identified four new gene loci associated with ABI that are also linked with coronary artery diseases (CAD) (6q26: *LPA* and 11q14.1: *DLG2*) or cholesterol levels (12q21.31: *TMTC2* and Xp21.1: *DMD*). Furthermore, we replicated a known ABI locus on cytoband 9p21.3 (*CDKN2B*) and four loci associated with PAD. In our TWAMA, we identified 145 blood transcripts associated with ABI at FDR 5% level. Gene set enrichment analysis of all TWAMA results revealed the inflammation-related pathways *interferon gamma response*, *neutrophil degranulation*, and *interferon alpha response* as the top three upregulated pathways in patients with lower ABI. Among overlapping genes between blood TWAMA and tissue-specific genetically regulated gene-expression association analysis, 24 genes showed consistent effect directions at nominal significance, with lower ABI-associated genes relating to stress response and vascular integrity, while higher ABI-associated genes linked to cellular homeostasis and metabolism.

**Conclusions:** In our integrated genome- and transcriptome-wide meta-analysis, we identified novel and confirmed known candidate genes and pathways associated with ABI. Association signals partly overlap with those of other cardiovascular traits such as CAD and carotid plaque formation. The integration of gene-expression data, validated known and added new molecular insight how inflammatory signaling can contribute to atherosclerosis and vascular dysfunction. These findings pave the way for improved understanding of the molecular underpinnings of PAD and inform future strategies for targeted prevention and therapy.

## Introduction

The Ankle Brachial Index (ABI) is an important non-invasive assessment of peripheral arteries and is used to screen for peripheral artery disease and general cardiovascular risk [1]. Normal ABI ranges from 1.00 to 1.40, ABI <0.90 is closely related to the incidence of Peripheral arterial disease (PAD) while ABI >1.40 indicates calcified or non-compressible vessels [1]. ABI has a high sensitivity in detecting PAD [2], while the specificity becomes lower in populations with certain conditions like Coronary Artery Disease (CAD) [3] or chronic kidney disease (CKD) [4]. Furthermore, ABI is also a predictor for the long-term cardiovascular health of a patient and can be used as first-line screening test for CAD because of its high specificity [5]. ABI values ≤0.90 are associated with an increased risk of major cardiovascular events, namely mortality, acute myocardial infarction, and ischemic stroke [6] and ABI values >1.40 are associated with an increased risk of all-cause mortality [7].

To better elucidate the molecular mechanisms affecting ABI, we integrate genome-wide genetic and transcriptomic association meta-analyses using measured or tissue-specific genetically inferred gene-expression profiles of two cohorts. Based on these findings, we perform pathway enrichment analysis to identify groups of genes associated with ABI and compare the findings of all three approaches on the gene and pathway level.

## Material and Methods

### Cohort description

We meta-analyse two studies collected in the LIFE Research Center for Civilization Diseases of the University of Leipzig, Germany. The LIFE-Adult study is a population-based cohort study of 10,000 participants from the city of Leipzig [8]. The majority of individuals in the study fall within the age range of 40 to 79, while a smaller subset of 400 participants are aged between 18 and 39. The study group consists of individuals of central European ancestry, and the primary aim of the study is to examine the prevalence, genetic predisposition, and influence of lifestyle-related factors (such as smoking, alcohol consumption, diet, and physical activity) on major chronic diseases, including subclinical markers of cardiovascular disease. The initial data collection for the study took place in between 2011 and 2014.

LIFE-Heart is an observational study of patients collected at the Heart Center of Leipzig, Germany, which is one of the worldwide leading Heart centers with a recruitment area covering several German federal states. The study design and a detailed description of patients can be found elsewhere [9]. A total of 6,994 patients were recruited with suspected or confirmed stable coronary artery disease (CAD) or myocardial infarction. Initial data collection was performed in between 2006 and 2014.

Baseline characteristics of both cohorts are provided at *Table 1*.

**Table 1:**
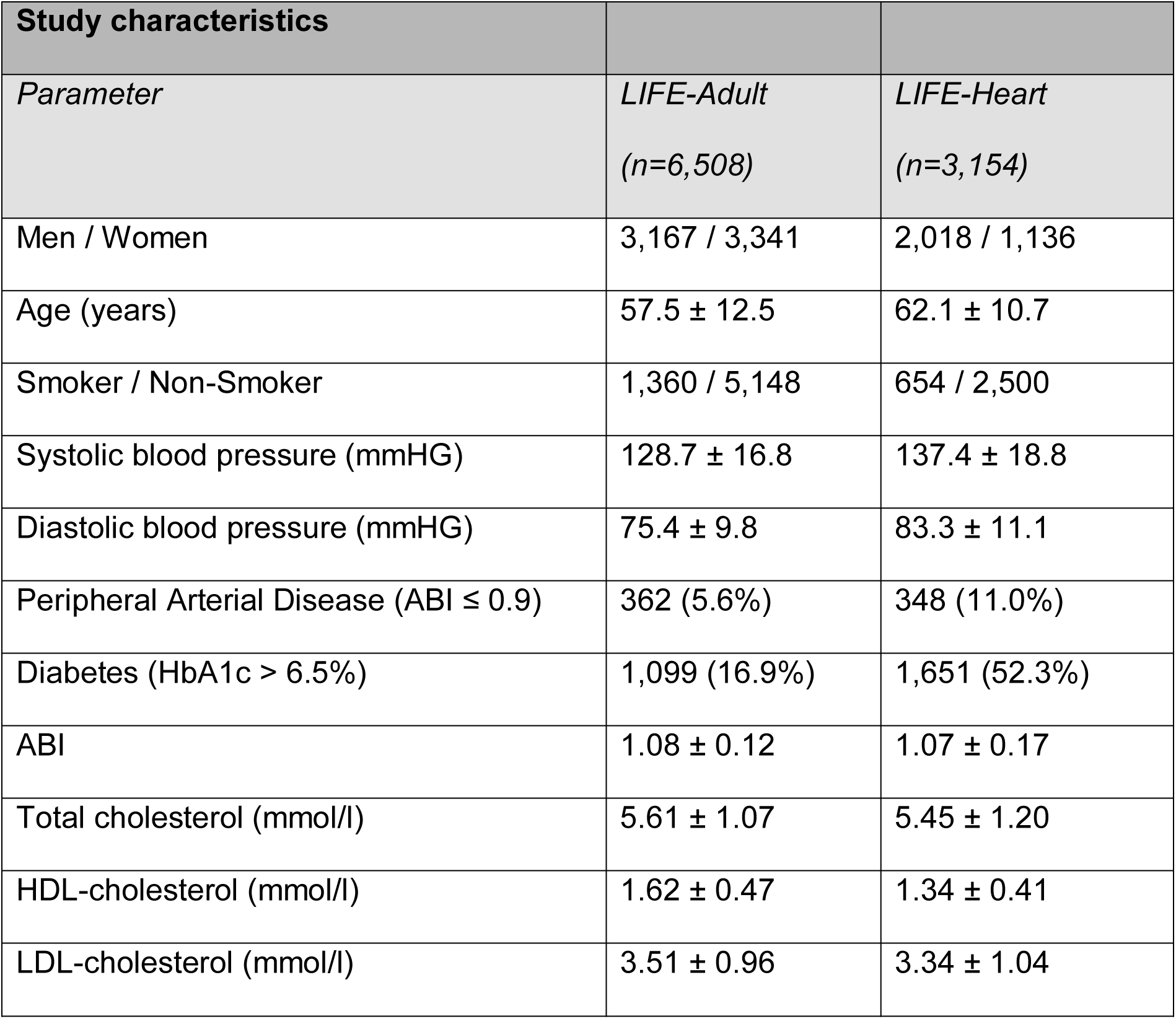
Study characteristics of LIFE-Adult and LIFE-Heart. Continuous parameters are presented as mean ± standard deviation. Analysis is restricted to participants / patients for which genotypes, ABI and smoking status are available. Former smokers and never smokers were summarized to non-smokers. Participants with ABI > 1.4 or on which percutaneous transluminal angioplasty had been performed were excluded.

| Study characteristics |  |  |
| --- | --- | --- |
| Parameter | LIFE-Adult<br>(n=6,508) | LIFE-Heart<br>(n=3,154) |
| Men / Women | 3,167 / 3,341 | 2,018 / 1,136 |
| Age (years) | 57.5 ± 12.5 | 62.1 ± 10.7 |
| Smoker / Non-Smoker | 1,360 / 5,148 | 654 / 2,500 |
| Systolic blood pressure (mmHG) | 128.7 ± 16.8 | 137.4 ± 18.8 |
| Diastolic blood pressure (mmHG) | 75.4 ± 9.8 | 83.3 ± 11.1 |
| Peripheral Arterial Disease (ABI ≤ 0.9) | 362 (5.6%) | 348 (11.0%) |
| Diabetes (HbA1c > 6.5%) | 1,099 (16.9%) | 1,651 (52.3%) |
| ABI | 1.08 ± 0.12 | 1.07 ± 0.17 |
| Total cholesterol (mmol/l) | 5.61 ± 1.07 | 5.45 ± 1.20 |
| HDL-cholesterol (mmol/l) | 1.62 ± 0.47 | 1.34 ± 0.41 |
| LDL-cholesterol (mmol/l) | 3.51 ± 0.96 | 3.34 ± 1.04 |

Both studies meet the ethical standards of the Declaration of Helsinki and were approved by the Ethics Committee of the Medical Faculty of the University Leipzig, Germany (LIFE-Adult: Reg. No 263-2009-14122009; LIFE-Heart: Reg. No. 276-2005). Written informed consent including agreement with molecular-genetic analyses was obtained from all participants.

### ABI assessment

ABI was determined taking into account the recommendation of the American Heart Association [10]. For LIFE-Adult, blood pressure was measured using oscillometry-, and photoplethysmography-based methods (Vicorder, Skidmore medical, UK) [11]. For both sides, we averaged the three available systolic blood pressure (SBP) measurements for each ankle and each upper-arm separately. We then calculated the ABI for the right side (left side) by dividing the average value for the right ankle (left ankle) by the higher value of the average values for the right and the left upper-arm. The lower of the two quotients was used as ABI assessment for the respective participant.

For LIFE-Heart blood pressure was measured by Doppler sonography sphygmomanometer cuffs and a hand-held Doppler probe (Huntleigh Mini-Dopplex, Germany) [9]. We averaged the two available SBP measurements for each ankle. For the upper-arms we only had two SBP measurements for the right side which we also averaged. ABI for the right side (left side) was calculated by dividing the average value for the right ankle (left ankle) by the average SBP of the upper-arm. The lower of the two results was considered as ABI for the respective participant.

For both cohorts, participants with ABI >1.4 were removed as ABI values above 1.4 indicate calcified vessels most common in patients with diabetes [12]. We also removed participants with percutaneous transluminal angioplasty or without genetic data.

### Genotyping

Both LIFE studies were genotyped using the Affymetrix Axiom SNP-array technology [13] (LIFE-Adult: CEU1 array, LIFE-Heart: CEU1 or CADLIFE array, a customized CEU1 array containing additional SNPs from CAD loci). For each study genotype calling was performed with Affymetrix Power Tools (v1.20.6 for LIFE-Adult CEU1; v1.17.0 for LIFE-Heart CADLIFE; v1.16.1 for LIFE-Heart CEU1), following best practice steps for quality control. These steps comprised filtering samples for signal contrast and sample-wise call rate, and included SNP filters in regards to platform specific cluster criteria. The LIFE-Heart datasets from the two distinct array platforms were combined after calling [14].

Samples with XY irregularities, including sex-mismatches or cryptic relatedness, and genetic outliers (>6 standard deviations of genetic principal components) were excluded. Further, variants with a call rate less than 0.97 and Hardy-Weinberg Equilibrium P < 1×10^-6^, were removed before imputation. Imputation was performed using the 1000 Genomes Project phase 3, Version 5, European reference panel [15] and IMPUTE2 [16] as software. In summary, 7,669 and 5,700 samples were successfully genotyped in LIFE-Adult and LIFE-Heart, respectively (7,660 and 5,688 samples for chromosome X). After filtering subjects with non-valid ABI measurement as described above, 6,508 and 3,154 individuals were included in our genetic association study, respectively.

### Gene Expression Analysis

For the LIFE-Adult study, whole blood was collected in Tempus Blood RNA Tubes (Life Technologies) and relocated to -80°C before RNA isolation. For the LIFE-Heart study, RNA was extracted from peripheral blood mononuclear cells (PBMCs, see Burkhardt et al. [17] and Holdt et al. [18] for a complete description of the sampling and measurement process).

Human HT-12.0 Version 4 Expression BeadChips (Illumina, San Diego, CA, USA) were used to measure gene expression in blood samples from the LIFE-Adult (whole blood) and the LIFE-Heart cohorts (PBMC). Both data sets were preprocessed separately using a similar procedure. Raw gene expression data was extracted using Illumina GenomeStudio without background correction. Expression values were log2-transformed and quantile-normalized [19]. Batch effects of expression BeadChips were corrected using the ComBat method [20]. The preprocessing was conducted in R / Bioconductor.

A total of 47,231 gene-expression probes are available on the array. Probes detected by Illumina GenomeStudio as expressed in less than 5% of the samples, showing associations with batch effects even after batch correction (with Bonferroni-adjusted p-value < 0.05) or without mapping to a gene according to IPA (ingenuity pathway analysis [21], were excluded. As a result, 36,374 (LIFE-Adult) and 36,368 (LIFE-Heart) gene-expression probes corresponding to 22,644 (22,641) genes remained in the data. From these probes, 36,366 corresponding to 22,640 genes were available in both data sets based on their probe IDs and were used in the meta-analysis.

With respect to gene-expression sample quality, we removed samples based on the following two conditions. First, the number of detected gene-expression probes of a sample had to be within ±3 interquartile ranges (IQR) from its median over all samples. Secondly, the Mahalanobis distance of several quality characteristics of a sample (signal of AmbionTM ERCC Spike-In control probes, signal of biotin-control-probes, signal of low-concentration control probes, signal of medium-concentration control probes, signal of mismatch control probes, signal of negative control probes and signal of perfect-match control probes) must not exceed the median + 3 IQR [22].

Overall, of the assayed 3,526 LIFE-Adult samples, 107 samples were excluded for quality reasons. Exclusion of the remaining duplicate samples (2 technical, 34 biological), samples that were not genotyped (189), samples with missing values of ABI, sex, age, smoking status, proportion of lymphocytes or proportion of monocytes (426), and patients with non-valid ABI measurement as described above (15) resulted in 2,753 samples available for further analysis.

Of the assayed 4,509 LIFE-Heart samples, 122 samples were excluded for quality reasons. Exclusion of duplicate samples (13), not genotyped samples (37), samples with missing values of ABI, sex, age, smoking status, proportion of lymphocytes and proportion of monocytes (1,766, of which 1,759 had missing ABI measurements), and patients with non-valid ABI measurement as described above (53), resulted in 2,518 samples ready for analysis.

### Statistical analysis

#### Genome-wide association meta-analysis (GWAMA)

We first performed genome-wide association analyses for ABI by linear regression models assuming an additive mode of inheritance with adjustments for sex, age and smoking status separately in each of the two LIFE studies using PLINK 2.0. We refrained from correction for population stratification due to low heterogeneity of study participants, and irrelevant association of population genetics principal components with the endpoint (supplementary table 1. Supplementary Figure 1). For LIFE-Adult (LIFE-Heart) only three (one) principal components were significantly correlated with a maximum explained variance of r²=1.6×10^-3^ for PC7 for LIFE-Adult (r²=2.7×10^-3^ for PC4 for LIFE-Heart).

All SNPs were harmonized between LIFE-Adult and LIFE-Heart to ensure the same effect allele. In addition, we checked for mismatching alleles or chromosomal position (hg19) with respect to 1,000 Genomes phase 3 European reference [15] and excluded SNPs with a high deviation of observed minor allele frequencies (absolute difference >20%). Only SNPs in the intersection of both studies were meta-analysed.

For genome-wide association meta-analysis (GWAMA), single study results per phenotype were combined using a fixed-effect model. We used I² statistics to evaluate heterogeneity and filtered findings with I²>90%. Only SNPs with samples-size weighted average minor allele frequency (MAF) ≥ 1%, and a samples-size weighted average minimum imputation info-score of at least 0.8, analyzed in both studies, were used.

The threshold for genome-wide significance was set to p<5×10^-8^. Associations with p<1×10^-6^ were considered as suggestive. In order to determine independent hits, variants, which are in LD r²≥0.1 with a SNP with stronger p-value, were considered tagged by that SNP (priority pruning). Pruned suggestive hits were assigned to loci by defining a region of ±500 kb around each SNP. In our hands, this resulted in non-overlapping regions.

A comprehensive annotation was applied to all SNPs that reached at least suggestive significant association levels using the following bioinformatics resources: Physically nearby genes were looked up from Ensemble [23] within ±250 kb around a variant. For associated variants, we assigned other GWAS hits in LD (r²>0.3) retrieved from the GWAS catalogue [24] and previously reported as well as in-house determined expression quantitative trait loci (eQTL) [24–32]. Combined Annotation-Dependent Depletion (CADD) score was used to assess variant deleteriousness [25].

Genome-wide summary statistics for GWAMA are available at zenodo [26].

#### Replication and look-up of known SNPs associated with atherosclerotic diseases

We looked up previously reported genetic loci for ABI, CAD, PAD and carotid plaque in the GWAS catalogue to replicate other reported ABI/PAD hits and to determine potential overlaps of genetic driving factors of ABI and other cardiovascular diseases as described in detail in Supplementary methods. For reported ABI and PAD hits, we also compared concordance of reported effect sizes with our data. Summary statistics of other studies were retrieved from the GWAS catalogue [27]. From these studies we selected SNPs with associations that showed at least suggestive significance (p<1×10^-6^). For each reported SNP, we identified all SNPs in our GWAMA ±1Mb of the reported SNP with LD r^2^≥0.8. In the following, we proceeded phenotype-wise for GWAS-catalogue studies reporting on phenotypes ABI, CAD, PAD and carotid plaque: When multiple GWAMA-SNPs were linked to the same GWAS-catalogue SNP, we selected the GWAMA-SNP with the highest linkage disequilibrium (prioritizing direct SNP match over higher LD based on R^2^ over shorter physical). When multiple association studies were reported for the same GWAS-catalogue SNP, we selected the GWAS-catalogue study with the highest evidence (prioritizing lower p-value over higher sample size). To avoid biased replication rates due to the LD structure in our data, we considered only a single assignment of each GWAS-catalogue SNP to a certain GWAMA SNP for all GWAMA SNPs tagged by the same variant in our priority pruning approach described above. Thereby, we prioritized those GWAS-catalogue SNP - GWAMA SNP pairs where the GWAS-catalogue studies had lower reported p-values over those linked with higher LD (based on R^2^) over those with shorter physical distance.

For the resulting SNP pairs, the effect allele and direction of effect were harmonized and compared between the literature and our meta-analysis results. We investigated replication at two levels: (1) a liberal criterion requiring nominal significance (one-sided p<0.05) with the expected effect direction (i.e., reverse effect directions for ABI compared with CAD, PAD or carotid plaque), and (2) a more stringent criterion requiring significance after Benjamini-Hochberg false discovery rate (FDR) correction at 5% level, maintaining the same directional requirements. Since we primarily aimed at validating reported ABI or PAD associations, we estimated the respective statistical power of our study in this regard. We assumed that the true effect was as reported in the GWAS catalogue. Standard errors and effective sample sizes were also retrieved from this resource. We also assumed that the observed effect (regression coefficient of the ABI in our data) was normally distributed and that the minor allele frequencies of each SNP in the original study and in our study were equal. For power calculation regarding the binary trait PAD, we divided the reported effects by π/√3 = 1.81 as recommended [28] and multiplied the reported standard errors by 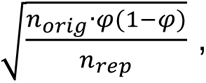 where φ denotes the proportion of PAD cases in the original study, and n_orig_ and n_rep_ denote the sample size in the original study and in our replication study, respectively [29].

#### Genetically-regulated gene-expression association analysis

We used the S-PrediXcan method [30] to associate genetically predicted gene-expression of four tissues with ABI: aorta, coronary and tibial artery as well as whole blood. S-PrediXcan is a computational method than can use summary statistics from GWAMA to predict gene expression levels of tissues based on genetic variants and to identify potential genetic associations of these inferred gene-expressions with other phenotypes. The method uses reference transcriptome data, which provides information on the expression levels of genes for different tissues, to construct prediction models of gene expressions based on genetic variants [31]. While we primarily analyse vascular tissue, blood was also analysed due to the superior expression quantitative trait loci (eQTL) models available for this tissue in GTEx [32]. Expression prediction models were downloaded from a respective GitHub repository [33] (see also PredictDB [34]). For data preparation, we lifted our data from hg19 to hg38 using the GWAS Summary Statistics harmonization tool [35]. Summary statistics of missing SNPs were imputed for the purpose of gene-expression prediction. We then tested the effects of gene expression variation on ABI in the four above mentioned tissues.

Genome-wide summary statistics for S-PrediXcan analysis are available at zenodo [26].

#### Transcriptome-wide association meta-analysis (TWAMA)

Gene-expression association analyses using directly measured blood transcriptomes were performed separately in the LIFE-Adult and LIFE-Heart cohort using limma [36]. Using the same linear model for both cohorts, each probe expression was treated as a response variable and ABI, sex, age, smoking status, proportion of lymphocytes, and proportion of monocytes as additive explanatory variables. In case lymphocyte and/or monocytes data were not available, these percentages were inferred using CIBERSORT [37] with percentage monocytes estimated as the difference between 100% and the sum of estimated neutrophils and lymphocytes. Results of single cohorts were again summarized by fixed-effect meta-analysis of the regression coefficients of ABI. Probes with I^2^ ≥ 50% were discarded and the p-values of the remaining probes were adjusted to control the false discovery rate (FDR) at 0.05 using the method of Benjamini and Hochberg [38]. Analyses were conducted in R version 4.1.1 [39].

Genome-wide summary statistics for TWAMA are available at zenodo [26].

#### Pathway analysis

We performed separate pathway analyses using the results of three gene expression analyses: 1) S-PrediXcan results of the artery tissue (aorta, coronary and tibial), 2) S-PrediXcan of the whole blood tissue, and, 3) TWAMA-results from differential expression of the directly measured blood gene expression data. In case of multiple transcripts per gene, transcripts were summarized to unique gene symbols by selecting the transcript with the smallest meta-analysis heterogeneity (Cochran’s Q) for each gene to enrich for associations homogeneous between cohorts. Fast pre-ranked Gene Set Enrichment Analysis (FGSEA) [40,41] of lists of ranked genes was performed using the R packages fgsea and clusterProfiler [42]. In two separate analyses, enrichment of Disease Ontology (DO) [43,44], and, according to previously published recommendations [45], of gene sets representing pathways culled from several sources were computed. Those included GO (biological process, excluding annotations that have evidence code IEA, i.e. inferred from electronic annotation, excluding ND, i.e. no biological data available, and excluding RCA, i.e. inferred from reviewed computational analysis), Reactome, Panther, NetPath, NCI, IOB, WikiPathways, MSigDB C2, MSigDB Hallmark and HumanCyc, available at http://download.baderlab.org/EM_Genesets/April_01_2022/Human/symbol/Human_GO_AllPathways_no_GO_iea_April_01_2022_symbol.gmt. For S-PrediXcan of the artery tissue, genes were ranked by their z-scores that had the highest absolute value across the three artery tissues (aorta, coronary, tibial). For S-PrediXcan of the whole blood tissue, genes were ranked by their z-scores. For directly measured gene expression, genes were ranked according to their z-statistics of the fixed-effect meta-analysis of the ABI regression coefficients (see Section “Transcription-wide association meta-analysis”). Gene sets of size between 15 and 500 were included in the enrichment analyses (1005 gene sets from the DO and 546 gene sets representing pathways). Gene sets with Benjamini-Hochberg FDR-adjusted p-value < 0.05 were considered significant.

## Results

### Genome-wide association meta-analysis (GWAMA)

After quality filtering, 8,828,968 SNPs remained for meta-analysis. No inflation of test statistic was observed (lambda = 0.9989, see Supplementary Figure 2 for a quantile-quantile plot). No hits were found at genome-wide significance level, but four loci showed suggestively significant associating SNPs (see Table 2 for an overview and Supplementary table 2 for more detailed association statistics, Fig 1 for a Manhattan Plot for ABI and Supplementary Figure 3 for regional association plots).

**Fig 1:**
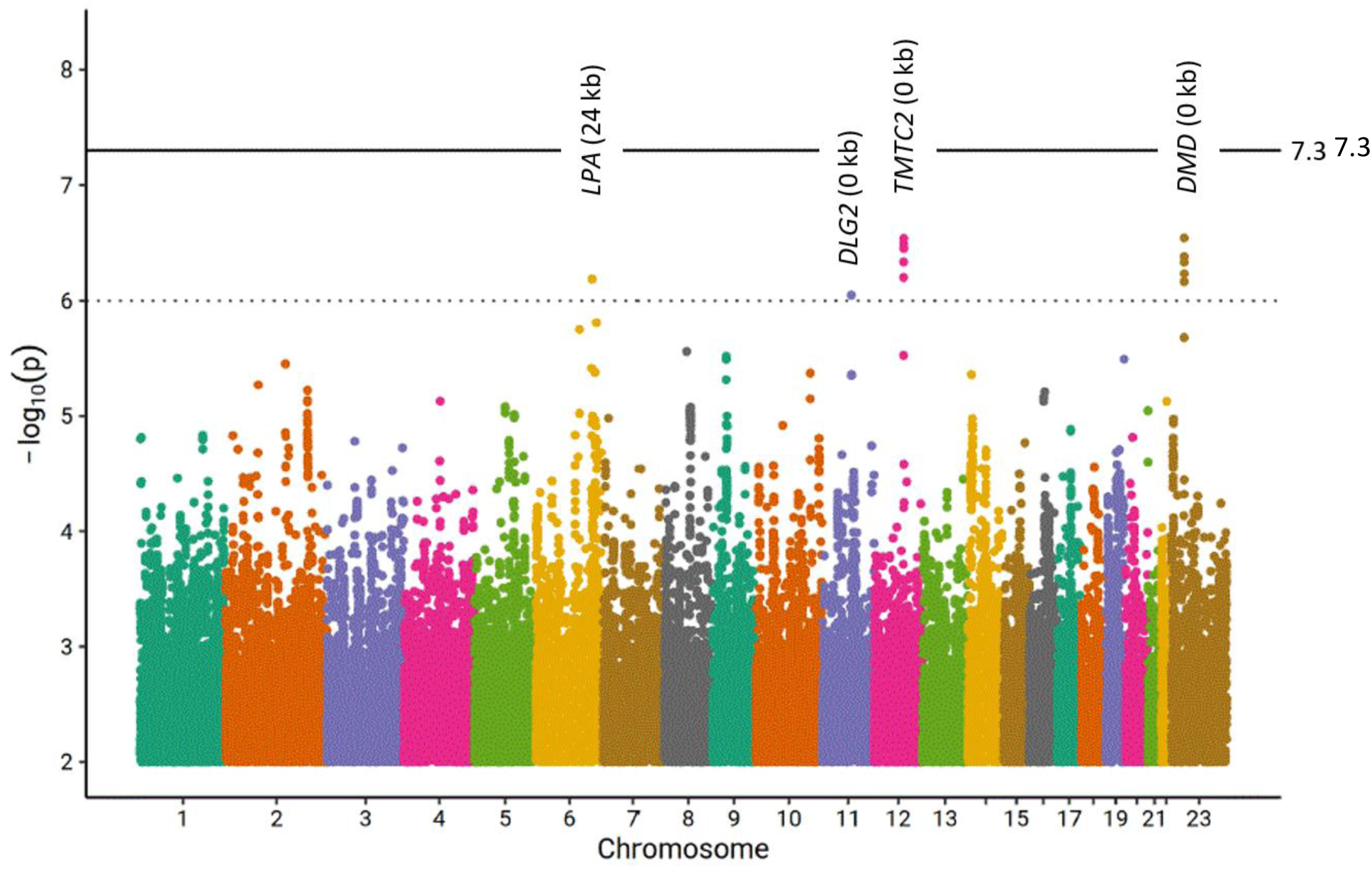
Manhattan plot of our genome-wide association analysis of ABI. Distribution of log10 transformed p-values. The bold line marks genome-wide significance (p<5×10^-8^). Four loci show signals of suggestive significance (p<1×10^-6^) but none of the signals achieved genome-wide significance. For each of these four loci, the nearest gene and the distance from the best associating SNP to this gene is reported

**Table 2:** Loci with suggestive evidence in our Meta-GWAS of ABI. We present the four loci with suggestive evidence, its respective top-SNPs and association statistics, candidate genes EA: effect allele, EAF: weighted average of effect allele frequency across studies, info score: weighted imputation info score across studies. I^2^: Heterogeneity estimate of meta-GWAS.

| Cytoband | Top-SNP | Candidate gene<br>(distance) | EA / other<br>allele | EAF | Info<br>score | $I^2$ | Beta (SE) | p-value |
| --- | --- | --- | --- | --- | --- | --- | --- | --- |
| 6q26 | rs186696265 | LPA (24 kb) | T / C | 0.02 | 0.94 | 0.61 | -0.358<br>(0.007) | $6.52 \times 10^{-7}$ |
| 11q14.1 | rs200811106 | DLG2 (0 kb) | TGG / T | 0.01 | 0.91 | 0 | -0.042<br>(0.008) | $8.98 \times 10^{-7}$ |
| 12q21.31 | rs80115038 | TMTC2 (0 kb) | T / C | 0.04 | 0.98 | 0 | -0.023<br>(0.005) | $2.90 \times 10^{-7}$ |
| Xp21.1 | rs5972515 | DMD (0 kb) | A / G | 0.05 | 0.98 | 0.57 | -0.018<br>(0.003) | $2.87 \times 10^{-7}$ |

We present the four loci with suggestive significant associations in more detail in the following. The lead SNP on cytoband 6q26 is rs186696265 (β = -0.036, p = 6.52×10^-7^) near *PLG* (12 kb) and *LPA* (24 kb). *LPA* is well known for controlling Lp(a) plasma concentrations also associating with a number of cardiovascular disease phenotypes including PAD [46]. The SNP was identified in GWAS studies of multiple endpoints including CAD, cholesterol and triglycerides (Supplementary table 3). The SNP was also in LD with several cis-eQTLs including the functionally plausible genes *ACAT2* involved in cholesterol metabolism [47], *SLC22A1* known for its role in elevated cholesterol and LDL-C levels [48] and *SOD2* linked to CAD [49]. Thus, this locus is functionally plausible but different candidate genes can be conceived.

On cytoband 11q14.1 the lead SNP is rs200811106 (β = -0.042, p = 8.98×10^-7^). This variant has a CADD score of 11.9 and is therefore likely to be deleterious. The SNP is in the coding sequence of *DLG2*, which has been associated with CAD before [50]. The SNP was also in linkage disequilibrium (LD) with variant rs142615018 in a GWAS associated with BMI (R^2^ =0.74, Supplementary table 3).

On cytoband 12q21.31 the top-associated variant rs80115038 (β = -0.023, p = 2.90×10^-7^) is in the coding sequence of *TMTC2*. *TMTC2* has been associated with HDL levels before [51]. No reported eQTL or associated GWAS were in LD with this variant.

The fourth locus is located at Xp21.1 with rs5972515 as lead SNP (β = -0.018, p = 2.87×10^-7^). This SNP is in the coding sequence of *DMD*. Dysregulation in *DMD* can cause Duchenne Muscular Dystrophy. Patients with this desease also show elevated cholesterol levels [52]. No reported eQTL or associated GWAS were identified. Thus, we could assign plausible candidate genes to all of our suggestive loci.

### Replication of reported associations and look-up of other cardiovascular trait associations

We looked-up four SNP sets that are reported for associations with ABI, PAD, CAD, and carotid plaque presence. Results are shown in supplementary table 5. A SNP association was considered successfully replicated, respectively co-associated with other cardiovascular disease traits, if it showed the expected effect direction (same for ABI, opposite for PAD, CAD, and Carotid plaque burden) and a one-sided p-value below 0.05 (liberal) or FDR<0.05 (stringent) (Table 3).

**Table 3:** Validation of reported associations and look-up of other CVD associations. For replication we compared our results with the unique published SNPs that had the lowest p-value across reported studies. We only consider data with identifiable effect direction. Details on replicated SNPs is provided in **Supplementary Table 5**, overlap of respective cytobands is shown in **Supplementary Figure 4 ^$^** one sided p-value <0.05. * significantly more than 5%, what would be expected by chance (binomial testing with p_expected_≤0.05).

| Reported trait association | Number of reported literature SNPs | Number of independent SNPs analysed | N SNPs replication level $p<0.05^{\$}$ | N SNPs replication level FDR<0.05 |
| --- | --- | --- | --- | --- |
| ABI | 14 | 6 | 1 (16.67%) | 1 (rs10757269 - CDKN2B) |
| PAD | 149 | 24 | 4 (16.67%)* | 0 |
| CAD | 2,557 | 342 | 28 (8.19%)* | 2 (rs186696265, intergenic; rs896655, intergenic) |
| Plaque | 52 | 13 | 2 (15.38%) | 1 (rs9644862, CDKN2B-AS1, CDKN2B) |

Of the 14 unique SNPs reported for association with ABI, six were reported with effect allele and beta estimate and corresponded to unique SNPs or proxys in our replication analysis. Of these, all had power above 80% (see **Supplementary Table 5** for details) and we were able to validate rs10757269 (9p21.3), which also showed an FDR below 0.05. The 9p21.3 locus is a well-known CAD locus, linked to regulation of cell proliferation [53].

Of the 149 unique SNPs reported for association with PAD, 24 were reported with effect allele and beta estimate allowing our validation analysis. Of these, 16 had power above 80% and we were able to validate four of them at nominal level. Rs118039278 (6q25.3, *LPA* locus), rs1537372 (again 9p21.3, *CDKN2B-AS1* locus), rs4722172 (7p15.3, *IL6*-locus) and rs6025 (1q24.2, Factor V Leiden locus) are all PAD-associated SNPs previously reported in a large population study (the Million Veteran Program with replication in the UK Biobank, [54]). However, none of these associations achieved an FDR below 0.05.

Our analysis of 342 independent CAD-associated SNPs revealed nominal significant ABI associations for 28 variants, exceeding what would be expected by chance (p_Enrich_ = 0.0079). Two of these SNPs achieved significant association at FDR ≤ 0.05: the intergenic variants rs186696265 (6q26) and rs896655 (again 9p21.3). Both variants were previously identified in a large-scale cardiovascular study, the UK Biobank with replication in CARDIoGRAMplusC4D [55]. The 6q26 locus (*LPA*) was also found in our ABI meta-GWAS.

In our examination of carotid plaque-associated variants (using an expanded phenotype definition detailed in the supplement), we found nominal ABI associations for two out of 13 independent SNPs. These included rs9644862 at the established risk locus 9p21 and rs1528152 at 7p21, with the latter having previously shown only suggestive evidence in the reporting study (p_original study_ = 3×10^-7^) [56]. The 9p21 association remained significant at FDR ≤ 0.05. Notably, 9p21 emerged as the only locus showing association evidence across all four analysed phenotypes: ABI, PAD, CAD, and carotid plaque underlining its high relevance for atherosclerosis development (**Supplementary Figure 4**).

### Transcriptome-wide association meta-analysis (TWAMA)

Next, we investigated genome-wide gene expression association in a TWAMA including measured gene expression data from the LIFE-Adult- and LIFE-Heart cohort. This identified 152 transcripts, representing 145 genes, showing significant (FDR < 0.05) positive (93) or negative (59) association with ABI. Statistics of significant associations are shown in supplementary table 6 and in Fig 2. For example, two probes of ALDH1A1 were among the top associations showing the strongest positive correlation with ABI.

**Fig 2:**
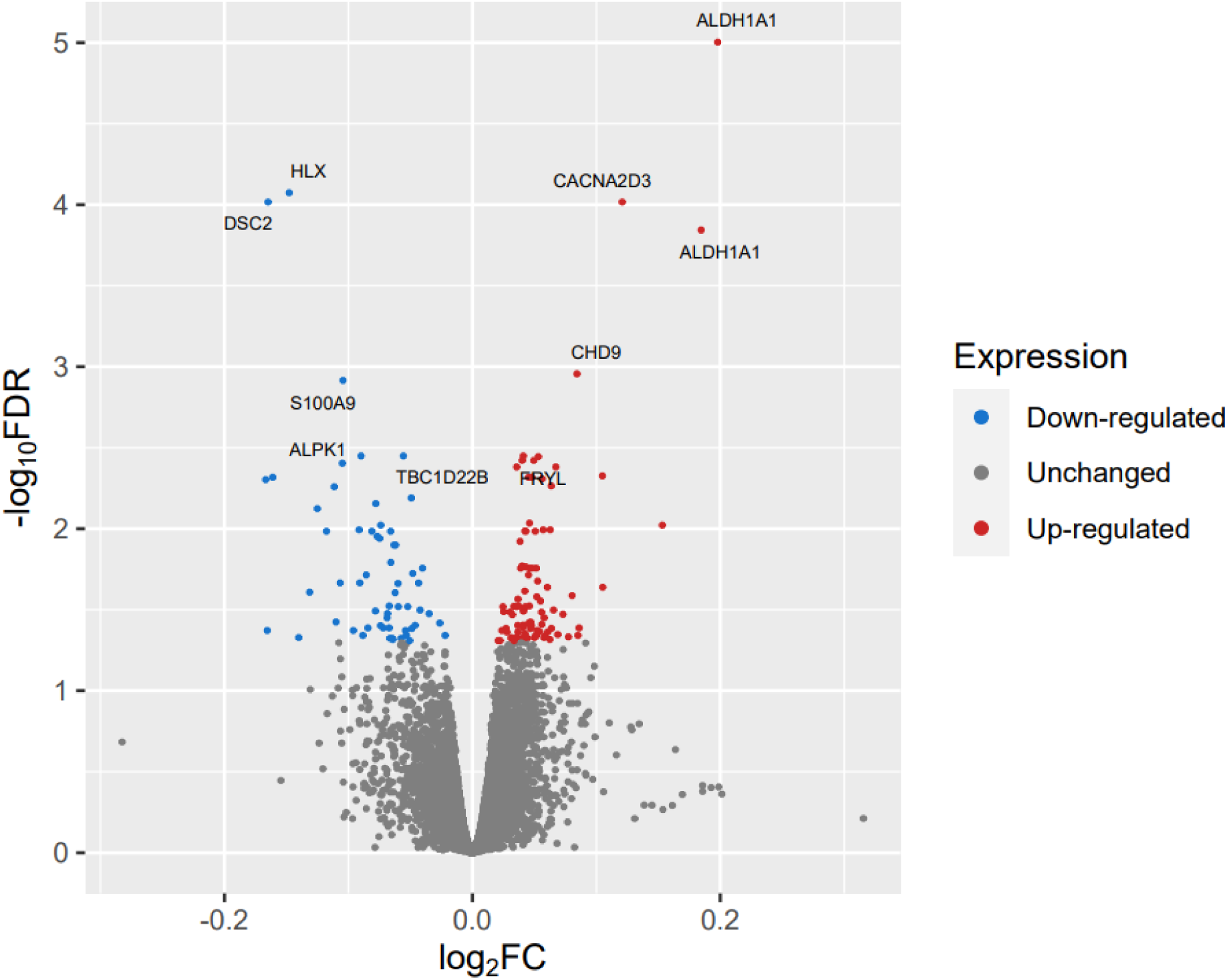
Volcano plot showing the association between measured gene-expression in blood and ABI. Fold changes and adjusted p-values were computed in the fixed-effect meta-analysis of the LIFE-Adult- and LIFE-Heart cohorts. We assigned gene names to the top ten associations.

Fast Gene Set Enrichment Analysis (FGSEA) applied to the results of the TWAMA identified 3 respectively 46 Disease Ontology terms (DOSE Ontology) as significantly (with adjusted p-value < 0.05) associated with an increase respectively decrease in ABI. Among the latter were terms as “atherosclerosis”, “arteriosclerotic cardiovascular disease”, “congestive heart failure” and “blood coagulation disease” (supplementary table 7, Figure 3A). In addition, we performed FGSEA of a comprehensive collection of gene sets representing various signalling pathways (Figure 3B). This analysis yielded 371 and 175 gene sets showing positive and negative correlation with the ABI, respectively (supplementary table 8). Gene sets most strongly associated with a decrease of the ABI were related to interferon-signalling, e.g., hallmark interferon gamma response (Figure 4).

**Fig 3:**
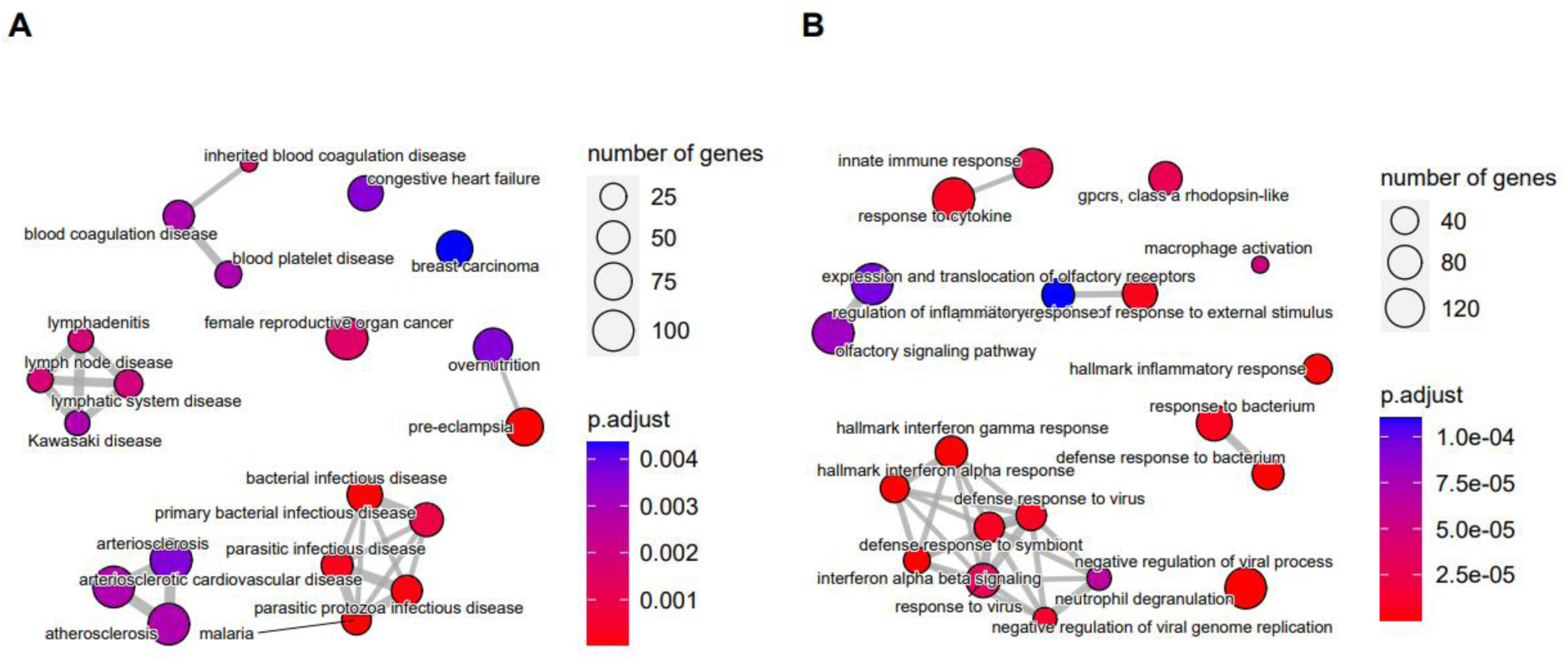
Enrichment plot of the top 20 significantly associating gene sets showing negative association with the ABI (i.e. increased expression in patients with PAD) from genome-wide results of TWAMA. A) Gene sets from the Disease Ontology, (B) gene sets collated from several databases of pathways. The placement of the gene sets and thickness of the connecting lines reflects the degree of similarity of the gene sets, i.e., number of common genes.

**Fig 4:**
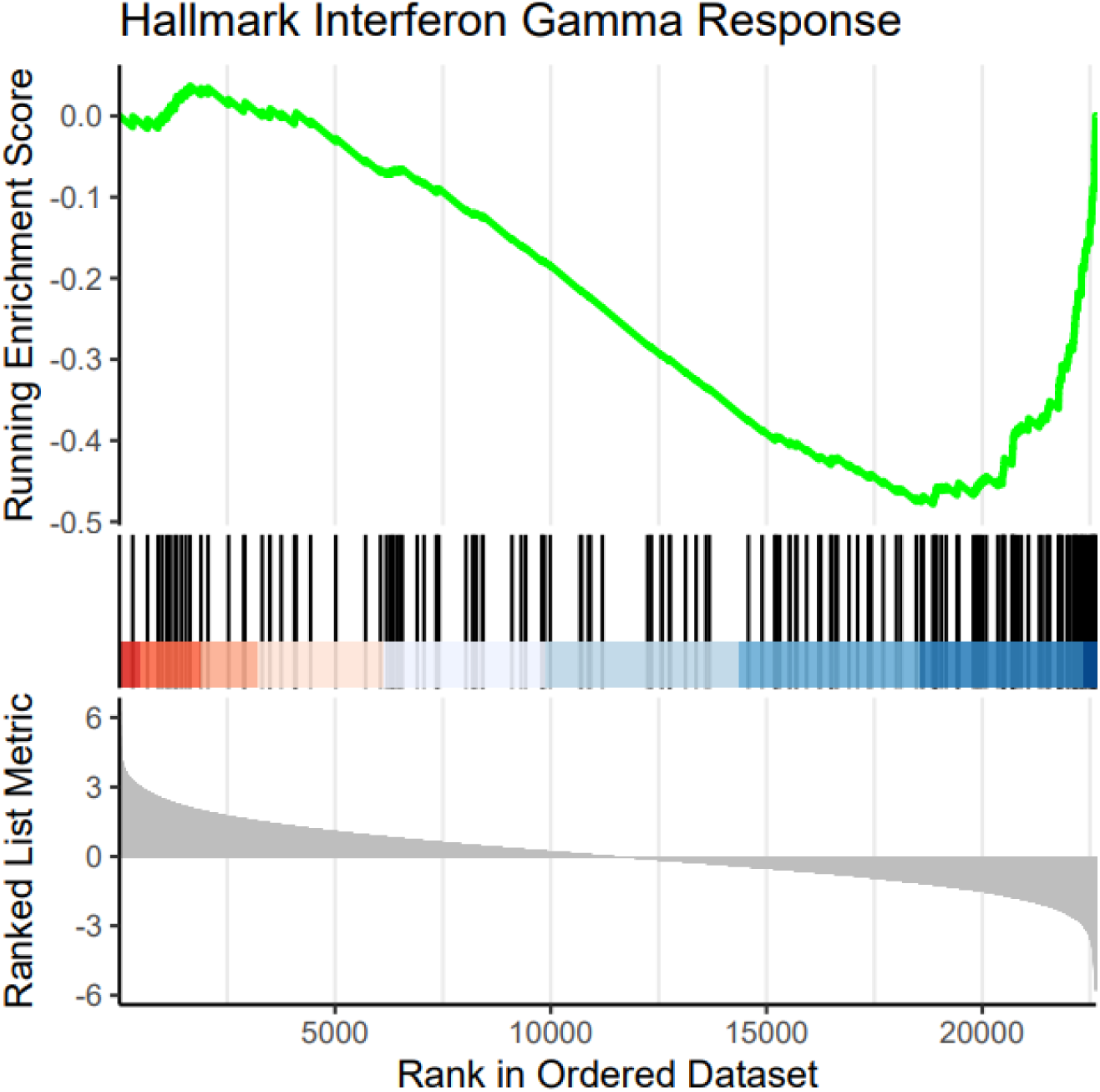
Gene Set Enrichment plot of the gene set representing the interferon gamma response pathway. This gene set is most strongly enriched at the bottom of the list of genes ranked by their association with the ABI, i.e., shows a strong negative association with the ABI (Benjamini-Hochberg adjusted p = 1.4×10^-8,^ Normalized Enrichment Score NES= -2.4).

### Genetically regulated gene-expression association analysis

Based on genetically controlled gene-expression following the S-PrediXcan framework, we identified 832 gene-expressions of artery tissues (tibial, aorta or coronary) and 586 gene-expressions of whole blood to be associated with ABI with nominal significance p-value < 0.05 in S-PrediXcan. However, none of these associations achieved an FDR value < 0.05. FGSEA analysis of association results from arterial tissues and whole blood did not reveal significant enrichment of gene sets from either Disease Ontology or signalling pathway collections (**Supplementary Table 9)**.

Nevertheless, when we stratified results of S-PrediXcan analysis to genes positively and negatively correlating with ABI, we observed a strong and highly significant enrichment of nominally significant associating genes overlapping between artery tissues and whole blood (OR 20.0, p_Fisher exact_=7.4×10⁻⁵⁶ for positively correlating genes; and OR 23.5, p_Fisher exact_=9.8×10⁻⁶⁴ for negatively correlated genes), indicating strong cross-tissue concordance of nominally significant associations. Identified overlapping genes are shown in **Supplementary Table 4**.

We then combined these results with our findings from TWAMA. **Fig 5** shows the overlap of associated genes identified in the three gene expression analyses: 1) tissue-based gene expression analysis of the arterial tissue (S-PrediXcan), 2) tissue-based gene expression analysis of the whole blood (S-PrediXcan), and 3) in the transcriptome-wide association meta-analysis of directly measured gene expression. We identified 6 consistently negative and 18 consistently positive correlated genes with ABI at nominal significance level in all three analyses (**Table 4**, all genes are shown in **Supplementary Table 10**).

**Fig 5:**
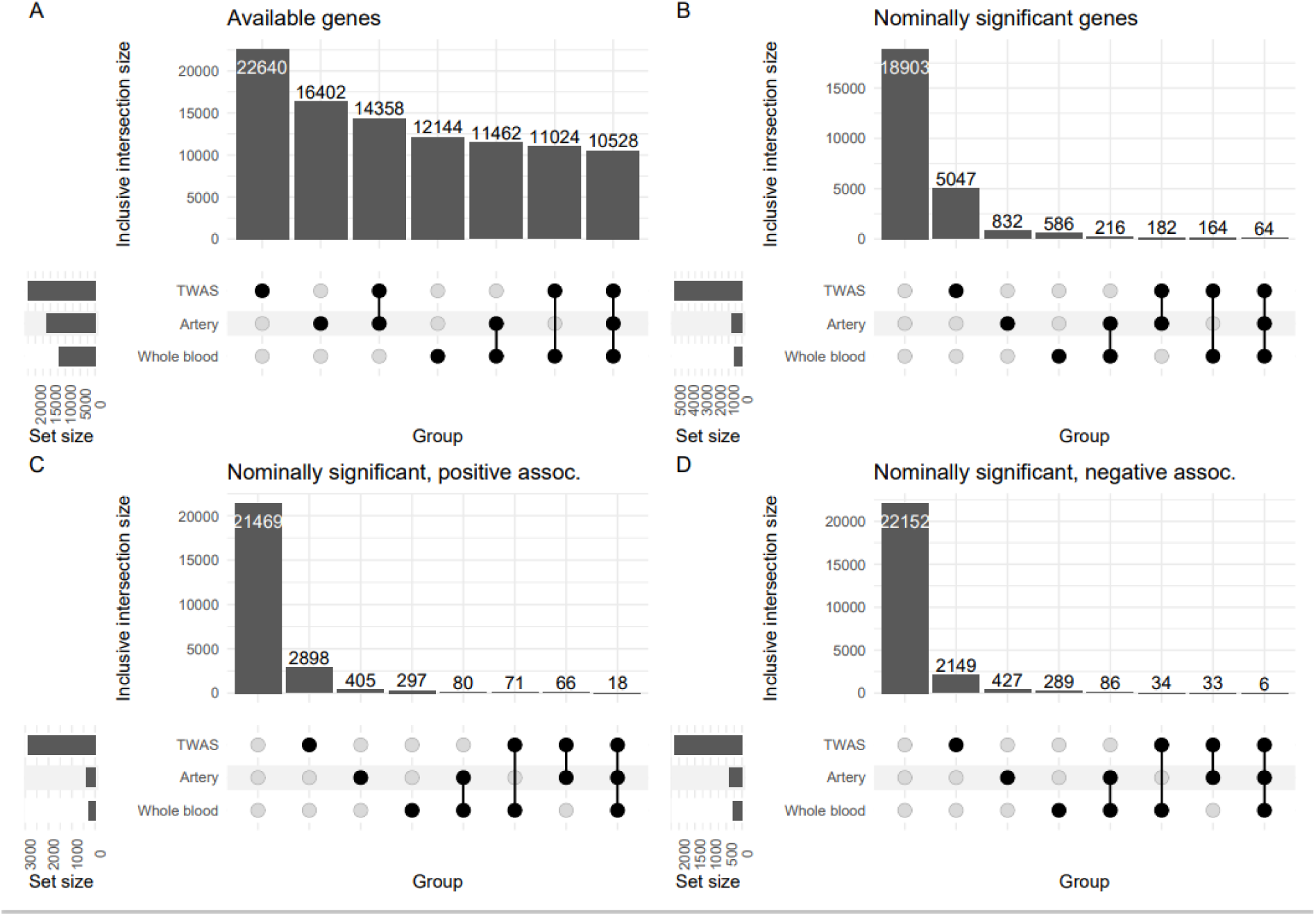
Upset plots showing numbers of genes that meet specific conditions in our three gene expression analyses: 1) tissue-based gene expression analysis of the arterial tissue, 2) tissue-based gene expression analysis of whole blood, and 3) transcriptome-wide association meta-analysis (TWAMA). A) numbers of available genes. B) numbers of nominally significant associating genes. C) and D) numbers of nominally significant associating genes showing a positive or negative association with the ABI, respectively. The shown intersections are inclusive, i.e., genes that belong to the sets defining an intersection may overlap with other sets if available. Details of overlapping genes is shown in **Supplementary Table 10**.

**Table 4:** Overlapping of nominally associated genes in blood TWAMA, blood S-PrediXcan and artery tissue S-PrediXcan analysis with consistent effect size. A total of 24 genes showed nominally significant associations in TWAMA as well as in artery tissue and whole blood. Overlap of all analyzed genes is shown in **Supplementary Table 10**

| Gene symbol | Z-score from arterial tissue S-PrediXcan | P-value from arterial tissue S-PrediXcan | Z-score from whole blood S-PrediXcan | P-value from whole blood S-PrediXcan | Z-score from measured expression TWAMA | P-value from measured expression TWAMA |
| --- | --- | --- | --- | --- | --- | --- |
| NDUFB6 | -2.43 | 3.2x10<sup>-02</sup> | -2.22 | 2.7x10<sup>-02</sup> | -3.04 | 2.3x10<sup>-03</sup> |
| PPP1R15A | -2.73 | 4.0x10<sup>-02</sup> | -2.73 | 6.4x10<sup>-03</sup> | -2.44 | 1.5x10<sup>-02</sup> |
| TGM2 | -2.25 | 2.4x10<sup>-02</sup> | -2.25 | 2.4x10<sup>-02</sup> | -2.38 | 1.7x10<sup>-02</sup> |
| PCNX1 | -3.27 | 2.4x10<sup>-03</sup> | -2.07 | 3.9x10<sup>-02</sup> | -2.21 | 2.7x10<sup>-02</sup> |
| GCLM | -2.46 | 4.9x10<sup>-02</sup> | -2.38 | 1.7x10<sup>-02</sup> | -2.13 | 3.3x10<sup>-02</sup> |
| CCM2 | -2.75 | 2.7x10<sup>-02</sup> | -2.40 | 1.6x10<sup>-02</sup> | -2.05 | 4.0x10<sup>-02</sup> |
| GOLT1B | 2.99 | 1.1x10<sup>-02</sup> | 2.25 | 2.4x10<sup>-02</sup> | 1.96 | 5.0x10<sup>-02</sup> |
| KDELC2 | 1.97 | 4.9x10<sup>-02</sup> | 1.97 | 4.9x10<sup>-02</sup> | 1.97 | 4.9x10<sup>-02</sup> |
| GALNT11 | 2.26 | 2.9x10<sup>-02</sup> | 2.26 | 2.4x10<sup>-02</sup> | 2.06 | 3.9x10<sup>-02</sup> |
| COPS6 | 2.37 | 5.0x10<sup>-02</sup> | 2.37 | 1.8x10<sup>-02</sup> | 2.09 | 3.7x10<sup>-02</sup> |
| RETSAT | 3.30 | 1.0x10<sup>-02</sup> | 2.89 | 3.8x10<sup>-03</sup> | 2.12 | 3.4x10<sup>-02</sup> |
| TXNDC16 | 3.20 | 1.4x10<sup>-03</sup> | 3.22 | 1.3x10<sup>-03</sup> | 2.13 | 3.3x10<sup>-02</sup> |
| PRRC1 | 2.83 | 4.1x10<sup>-03</sup> | 2.34 | 1.9x10<sup>-02</sup> | 2.18 | 3.0x10<sup>-02</sup> |
| CYTH1 | 2.47 | 3.9x10<sup>-02</sup> | 2.47 | 1.4x10<sup>-02</sup> | 2.20 | 2.8x10<sup>-02</sup> |
| ALDH7A1 | 1.93 | 4.8x10<sup>-02</sup> | 2.37 | 1.8x10<sup>-02</sup> | 2.33 | 2.0x10<sup>-02</sup> |
| LRRC23 | 2.17 | 3.1x10<sup>-02</sup> | 2.01 | 4.5x10<sup>-02</sup> | 2.37 | 1.8x10<sup>-02</sup> |
| PKIG | 2.51 | 1.3x10<sup>-02</sup> | 2.51 | 1.2x10<sup>-02</sup> | 2.60 | 9.2x10<sup>-03</sup> |
| CEP192 | 2.32 | 2.4x10<sup>-02</sup> | 2.22 | 2.6x10<sup>-02</sup> | 2.61 | 8.9x10<sup>-03</sup> |
| GGACT | 2.44 | 2.7x10<sup>-02</sup> | 2.19 | 2.8x10<sup>-02</sup> | 2.67 | 7.5x10<sup>-03</sup> |
| NIFK | 2.36 | 1.8x10<sup>-02</sup> | 2.36 | 1.8x10<sup>-02</sup> | 3.03 | 2.4x10<sup>-03</sup> |
| SNX16 | 2.50 | 4.2x10<sup>-02</sup> | 2.30 | 2.1x10<sup>-02</sup> | 3.15 | 1.6x10<sup>-03</sup> |
| TRMT61B | 2.64 | 8.8x10<sup>-03</sup> | 2.64 | 8.3x10<sup>-03</sup> | 3.37 | 7.5x10<sup>-04</sup> |
| RABL2B | 2.67 | 9.5x10<sup>-03</sup> | 2.54 | 1.1x10<sup>-02</sup> | 4.05 | 5.1x10<sup>-05</sup> |
| RAD51C | 3.14 | 7.1x10<sup>-03</sup> | 3.14 | 1.7x10<sup>-03</sup> | 4.17 | 3.1x10<sup>-05</sup> |

The genes negatively correlated with ABI in all three analyses prominently feature in cellular stress response and vascular integrity pathways. Key examples include *GCLM*, central to glutathione synthesis and cellular antioxidant defence [57], and *PPP1R15A* (*GADD34*), which regulates the integrated stress response [58]. CCM2 is crucial for endothelial stability and vascular integrity [59]. *TGM2*’s involvement in vascular stiffness and inflammation [60] further underscores the connection between these genes and vascular health.

In contrast, the genes positively correlated with ABI primarily cluster around pathways essential for protein homeostasis, cellular trafficking, and metabolic regulation. This group includes genes involved in protein processing and transport (*GOLT1B*, *KDELC2*), protein glycosylation (*GALNT11*), and cellular quality control (*COPS6* as subunit of the *COP9* signalosome complex). Additionally, *RETSAT* plays a role in retinol metabolism, which is significant for vascular health due to vitamin A’s role in cell differentiation [61]. Several genes in this group also contribute to cellular integrity, such as *ALDH7A1* in cellular detoxification and *RAD51C* in DNA repair and genomic stability.

## Discussion

In our study, we performed a comprehensive integrative analysis of genetic and transcriptomic factors influencing ABI. The results contribute to the understanding of the genetic architecture of ABI and identify a multitude of potential new genes effecting vascular health.

In our genetic meta-analysis of two cohorts no significant inflation of the test statistic was observed, indicating reliable results and efficient data pre-processing. While no genome-wide significant associations were found, we identified four suggestive loci to which we can assign biologically plausible candidate genes. In our TWAMA one probe of *DMD* showed nominal significance (p=0.046). The association with CAD relevant loci (*LPA* and *DLG2*) needs further evaluation to better understand the underlying mechanisms of action. For *TMTC2* and *DMD* the cause and effect is well understood due to their involvement in cholesterol metabolism. Cholesterol levels are a major contributing factor to cardiovascular disease [62,63].

We furthermore validated previously reported SNPs for their associations with ABI, PAD, CAD, and carotid plaque burden. While we were not able to identify SNPs with genome-wide significance for ABI in our data, we replicated one ABI and four PAD SNPs and identified co-association of 28 CAD and two plaque SNPs at nominal level. These are more associations at this level than expected by chance, supporting genetic commonalities between these different vascular endpoints.

Measured blood gene-expression association analysis identified 145 differentially expressed genes at FDR 5% level. The fact that we identified more direct associations with measured gene-expression than with analysis of genetic regulated tissue specific expression (S-PrediXcan) suggests that context specific gene-expression is more relevant than genetically regulated gene-expression. Pathway analysis of all TWAMA genes resulted in plausible pathways and points towards interferon-signalling as a potential driving factor for PAD molecular pathology. The strong association between interferon gamma response pathways and ABI identified in our analysis is in line with existing research [64]. In a recent study, hallmark interferon gamma response was one of the most significantly enriched pathways in macrophages from symptomatic human atherosclerotic plaques [65].

We combined TWAMA results to integrate information from measured gene expression data with tissue-specific, genetic driven expression changes. Overlapping genes at nominal association significance were also plausible: Genes with expression levels correlating with lower ABI values were associated with increased activation of stress response pathways and compromised vascular integrity [57–60], while genes with expression levels correlating with higher ABI values were related to enhanced cellular homeostasis, efficient protein quality control mechanisms, and metabolic regulation [61] (**Table 4** and **Supplementary Table 10**). This underscores the value of the overlapping genes as candidate genes for pathomechanistical follow up-studies, to confirm that in vascular health, compromised circulation can involve cellular stress and altered protein homeostasis, whereas robust cellular function can support healthy vascular tissue.

We acknowledge several limitations of our study: 1) We investigated a limited sample size as our meta GWAS included a total of 9,662 people and lacked an independent replication cohort. However, we provided full summary statistics of each analyzed cohort enabling future meta-analyses to confirm suggestive loci identified by us. 2) We only considered European subjects, hence, generalisation of our findings to other ethnicities needs to be explored in future studies. 3) We could not validate identified top-genes in functional studies, however, we provide reported functional evidence where available.

In conclusion, while no genome-wide significant ABI-associated SNPs emerged, our integrative genetic and transcriptomic analyses revealed suggestive signals and identified genes whose expression correlates strongly with ABI. The enrichment of nominally significant overlapping genes in both arterial tissues (including the aorta) and whole blood, and their directional consistency with ABI, highlights systemic regulatory mechanisms underlying peripheral arterial health. Our findings point toward pathways involved in cellular stress response, vascular integrity, metabolic regulation, and interferon-signaling as potential contributors to ABI variation. These results provide a foundation for future investigations aiming to understand the molecular determinants of vascular function and peripheral arterial disease risk.

## Data Availability

Genome-wide summary statistics for GWAMA, S-PrediXcan and TWAMA are available at zenodo (https://doi.org/10.5281/zenodo.11526126).

https://zenodo.org/records/11526126

https://doi.org/10.5281/zenodo.11526126

## Acknowledgments

We thank the participants of LIFE-Adult and LIFE-Heart very much for their time and blood samples. We thank Kay Olischer and Annegret Unger very much for ABI assessment of the LIFE-Heart participants.

## Author contribution

Michael Rode: manuscript writing, data analysis

Maciej Rosolowski: contribution to data analysis, contribution to manuscript writing, supervision of analyses

Sylvia Henger: study management of LIFE-Heart, analysis of data quality

Kerstin Wirkner: study management of LIFE-Adult

Katrin Horn: data analysis

Andrej Teren: patient investigation

Markus Loeffler: PI of LIFE-Adult study

Joachim Thiery: PI of LIFE-Heart study

Janne Pott: contribution to data analysis

Holger Kirsten: contribution to manuscript writing, contribution to and supervision of analyses

Markus Scholz: study design, supervision, contribution to interpretation and manuscript writing

All authors red and approved the final manuscript.

## Supporting information

**Supplementary Figure 1:**
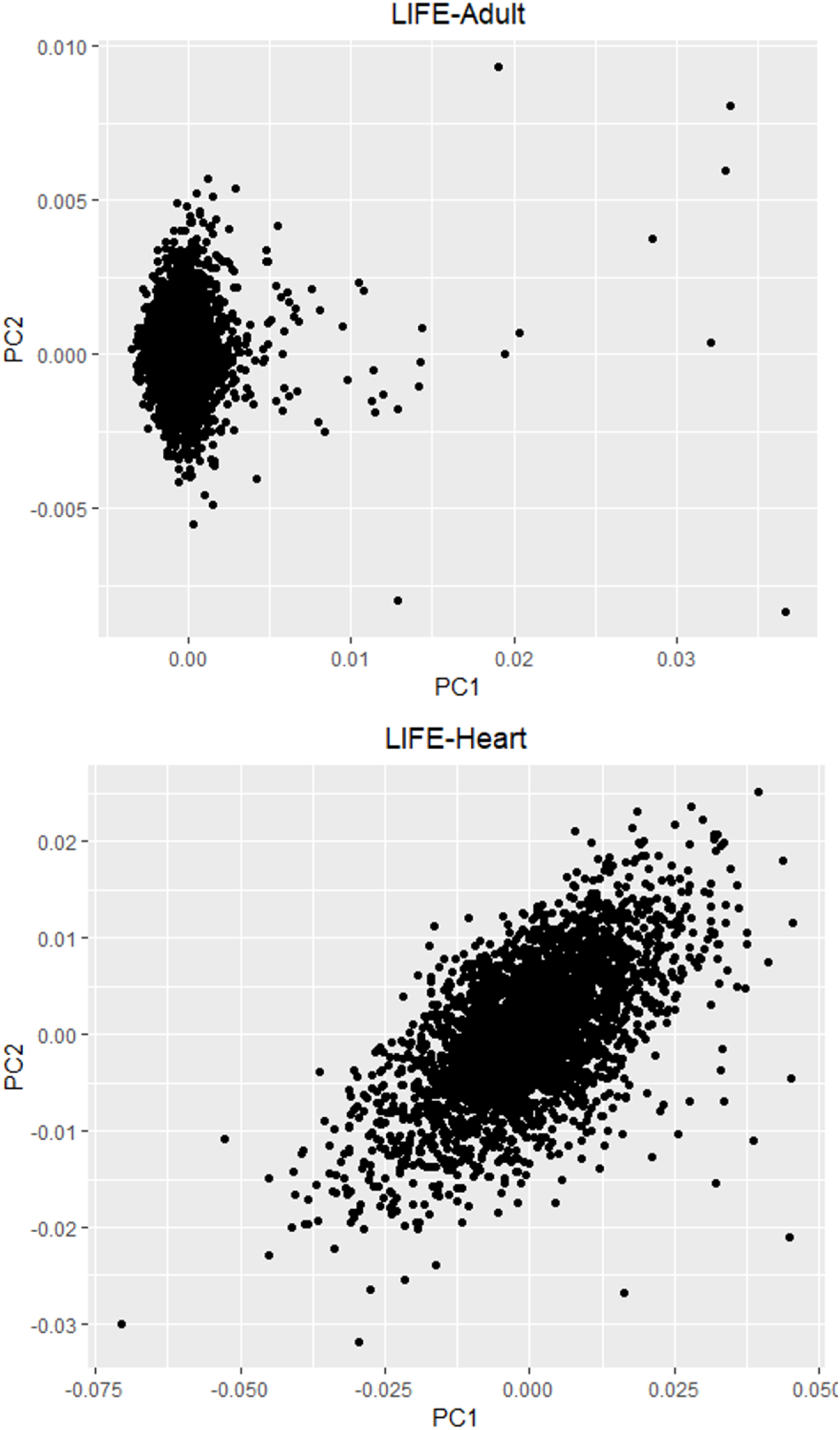
Principal component analysis of first two principal components for LIFE-Adult and LIFE-Heart

**Supplementary Figure 2:**
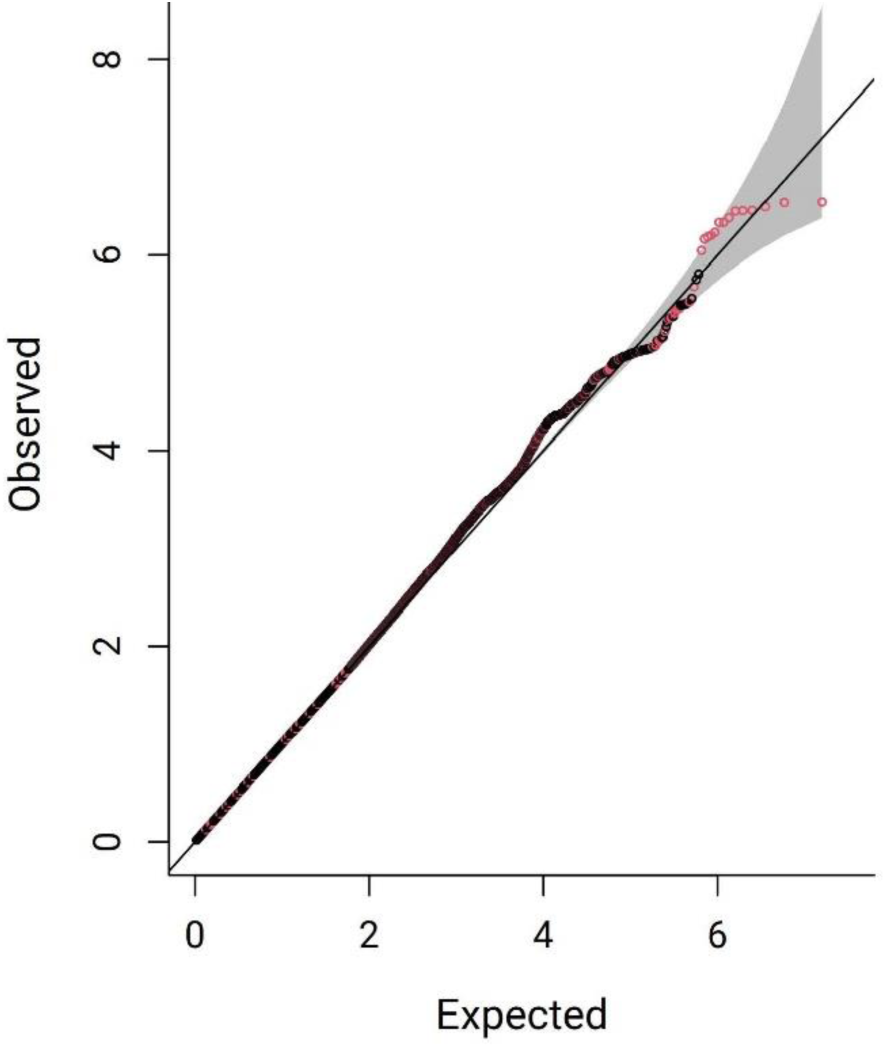
QQ Plot showing the observed quantiles vs the expected Chi-squared statistic. The factor lambda was 0.999, i.e. not indicating any evidence for inflation of test statistics. Red circles indicate SNPs with minor allele frequency < 0.05

**Supplementary Figure 3:**
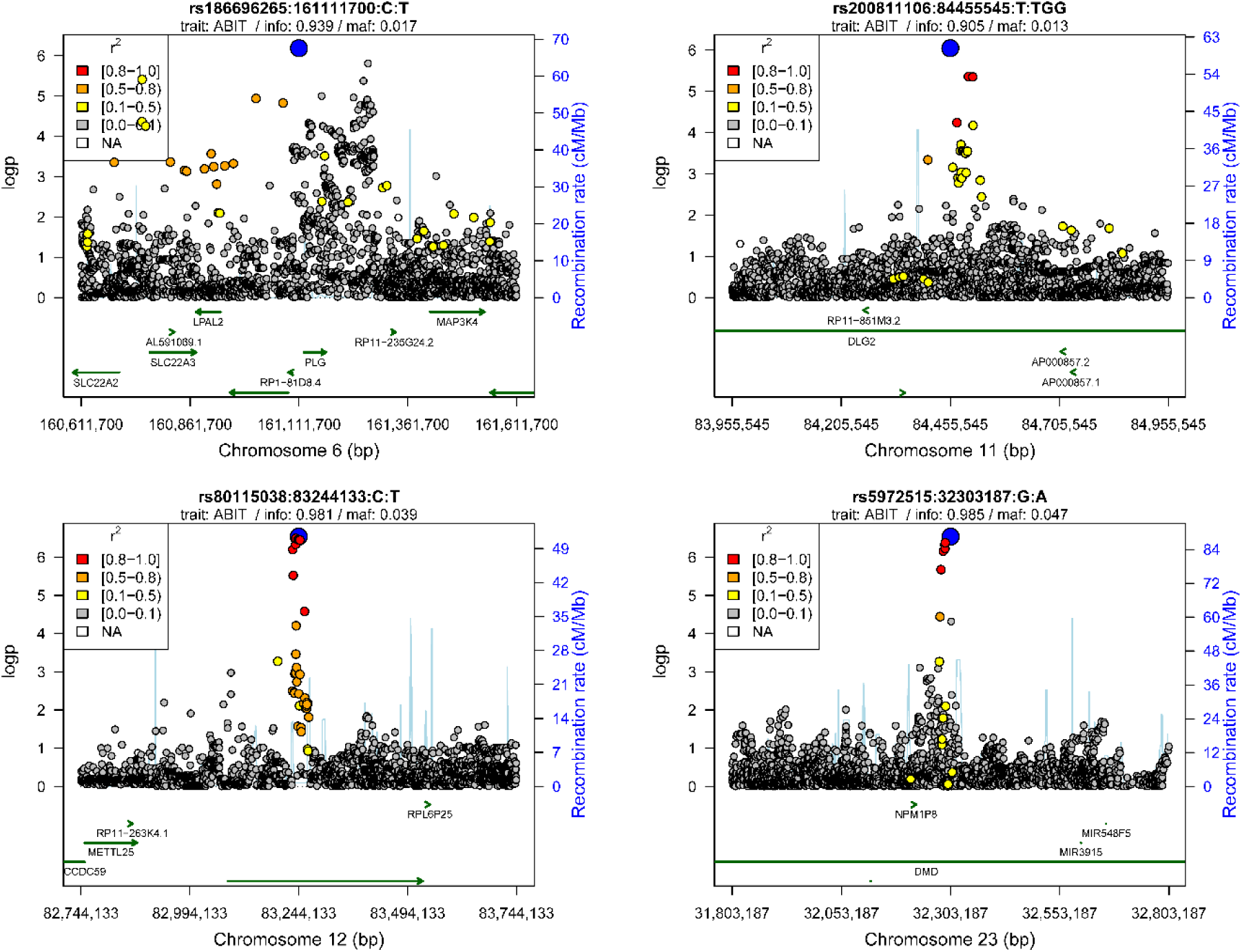
Regional ABI association plots for our four top SNPs. The top SNPs are coloured blue, the other SNPs according to their LD with the respective lead SNP). The top SNPs are always supported by additional variants.

**Supplementary Figure 4:**
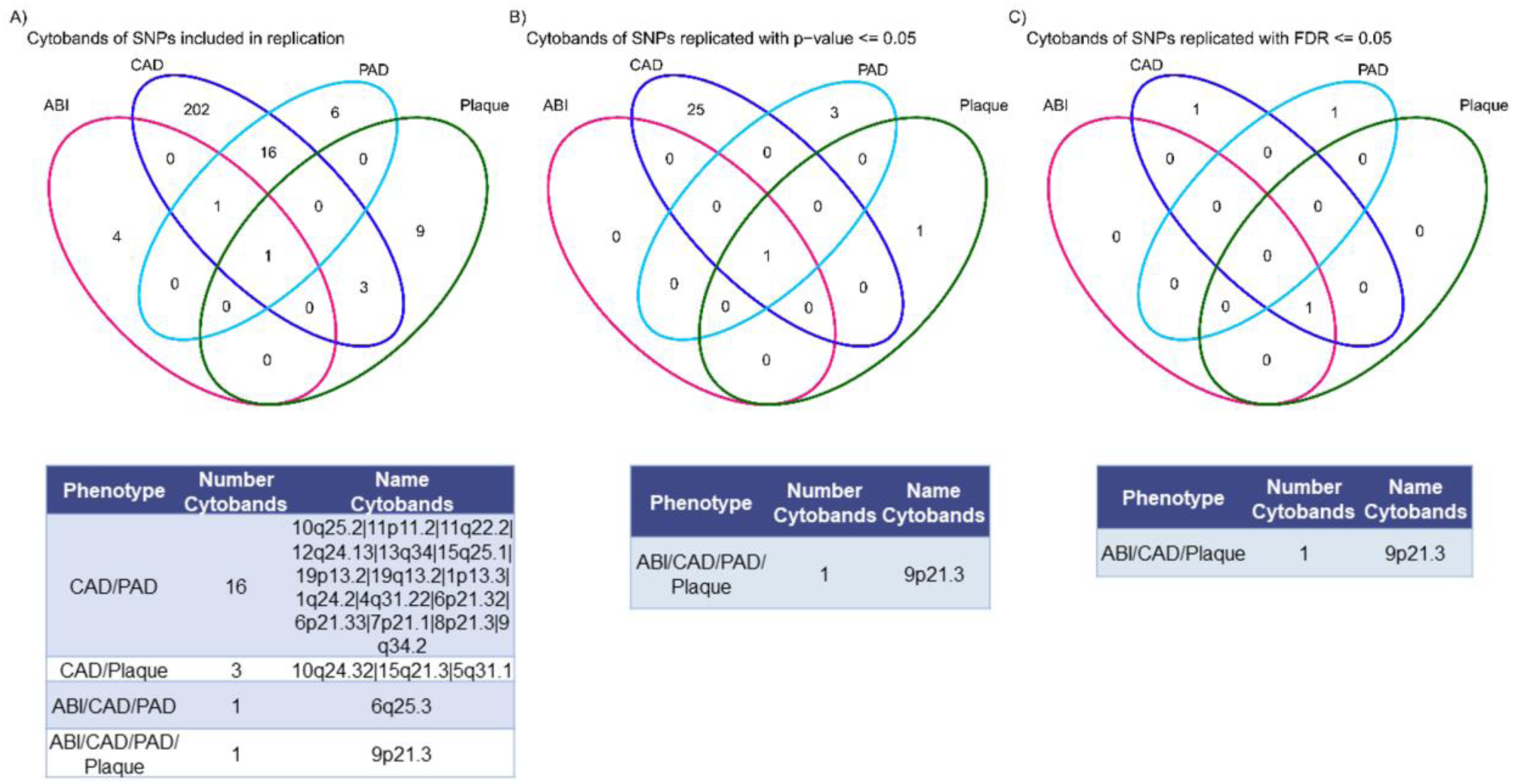
Overlap of loci included in replication/co-association analysis. **A)** All variants, **B)** Nominally significant variants, **C)** Associations significant at FDR 5%. The tables show associations overlapping between at least two traits.

