## Supplementary Methods for "Genome- and Transcriptome-wide association meta-analysis reveals new insights into genes affecting coronary and peripheral artery disease"

The GWAS catalogue was downloaded in 2022, Dec, 15, 16, 16, 21, for CAD, PAD, Plaque, and ABI-related phenotypes, respectively. Reported association with p-value > $10^{-6}$ were not considered.

Phenotypes included in carotid plaque burden:

Sum of carotid plaque area

Carotid plaque maximum area

Mean area of carotid plaque

Maximum stenosis

Sum of stenosis

References used to identify candidate genes are as follows:

ABI [1–4],

CAD [5–56],

PAD [2,3,19,46,57–62]
